# Genetically predicted on-statin LDL response is associated with higher intracerebral hemorrhage risk

**DOI:** 10.1101/2022.03.22.22272697

**Authors:** Ernst Mayerhofer, Rainer Malik, Livia Parodi, Stephen Burgess, Andreas Harloff, Martin Dichgans, Jonathan Rosand, Christopher D Anderson, Marios K Georgakis

## Abstract

Statins lower low-density lipoprotein (LDL) cholesterol and are widely used for the prevention of atherosclerotic cardiovascular disease. Whether statin-induced LDL reduction increases risk of intracerebral hemorrhage (ICH) has been debated for almost two decades. Here, we explored whether genetically predicted on-statin LDL response is associated with ICH risk using Mendelian Randomization. Utilizing genomic data from randomized trials, we derived a polygenic score from 35 single nucleotide polymorphisms (SNP) of on-statin LDL response and tested it in the population-based UK-Biobank (UKB). We extracted statin drug and dose information from primary care data on a subset of 225,195 UKB participants covering a period of 29 years. We validated the effects of the genetic score on longitudinal LDL measurements with generalized mixed models and explored associations with incident ICH using Cox regression analysis. Statins were prescribed at least once to 75,973 (31%) of the study participants (mean 57 years, 55% females). Among statin users, mean LDL decreased by 3.45 mg/dl per year (95% CI: [-3.47, -3.42]) over follow-up. A higher genetic score of statin response (one SD increment) was associated with significant additional reductions in LDL levels (−0.05 mg/dl per year, [-0.07, -0.02]), showed concordant lipidomic effects on other lipid traits as statin use, and was associated with a lower risk for incident myocardial infarction (HR per SD increment 0.98 95% CI [0.96, 0.99]) and peripheral artery disease (HR per SD increment 0.92 95% CI [0.86, 0.98]). Over a 11-year follow-up period, a higher genetically predicted statin response among statin users was associated with higher ICH risk in a model adjusting for statin dose (HR per SD increment 1.15, 95% CI [1.04, 1.27]). On the contrary, there was no association with ICH risk among statin non-users (*p*=0.89). These results provide further support for the hypothesis that statin-induced LDL reduction may be causally associated with ICH risk. While the net benefit of statins for preventing vascular disease is well-established, these results provide insights about the personalized response to statin intake and the role of pharmacologic LDL-lowering in the pathogenesis of ICH.

## Introduction

Intracerebral hemorrhage (ICH) is a devastating disease associated with a 50% 30-day mortality and major disability among survivors.^1,2^ HMG-CoA-reductase inhibitors, commonly known as statins, reduce low-density lipoprotein (LDL) and are widely used for prevention of atherosclerotic cardiovascular disease.^3^ The role of LDL in the pathogenesis of intracerebral hemorrhage (ICH) and whether statin intake increases ICH risk has been a matter of continued debate.^4,5^ Early clinical trials of statin administration had found a slightly elevated risk for ICH among statin users,^5-7^ which was in line with data from prospective observational studies demonstrating that increased serum total cholesterol and LDL levels are negatively associated with ICH risk in a dose-dependent manner.^8-13^ Although subsequent meta-analyses of statin trials found inconsistent results for overall statin use and risk of ICH,^12,14-17^ high-dose statin use remained associated with an increased ICH risk.^18^ However, post hoc analyses from statin trials could not detect statistically significant increases in ICH risk associated with aggressive LDL lowering to <70 mg/dl^19^ or <55 mg/dl.^20^ These conflicting data about incident ICH among statin users remain a source of concern among medical professionals and are the motivator of the ongoing NINDS-sponsored Statins in Intracerebral Hemorrhage (SATURN) randomized trial (NCT03936361).

Human genetic data are a valuable resource for unraveling the role of specific mechanisms in the pathogenesis of human disease and for identifying putative causal associations. Because genetic liability to polygenic traits is randomly assigned at birth, using genetic variants that are reliably associated with a trait of interest but do not vary with correlated confounders can reduce bias from confounding in associations between genetically predicted traits and outcomes.^21-23^ This framework, named Mendelian randomization (MR) has previously been widely used for establishing causal associations in the field of cerebrovascular disease.^24 25^Applying this concept in the association between statin use and ICH, previous studies have explored whether genetically predicted levels of lipids influence the risk of ICH.^26-29^ Genetic variants in the *CETP* gene, responsible for higher HDL cholesterol, were associated with an increased risk for ICH in case-control studies, ^27,28^ and genetically predicted lower LDL levels raised the risk for ICH among participants of the UK and China Kadoorie Biobanks.^26,29^ However, these findings do not address the specific question of whether statin-induced LDL-lowering actually increases ICH risk, because the genetic variants capture lifelong small effects on blood lipid levels and not the much stronger short-term effects that result from taking a medication in adulthood.

Expanding the MR concept, genetic variants associated with physiologic response to a drug could be used for stratifying individuals according to their innate sensitivity to it. Because genetic variants driving drug response are not known to physicians at the time of prescription, they could be used as instruments for randomizing participants of observational studies to different levels of exposure to the drug under study. This randomization could enable the determination of dose-dependent effects of specific drugs on potential side-effects or for exploring repurposing opportunities with the use of observational data, thus overcoming key limitations of conventional MR analyses.^30^ In this study, we applied this concept to study the relationship between statin use and ICH risk by leveraging large-scale genetic data for on-statin LDL response from clinical trials and population-based observational data from the UK Biobank.

## Materials and Methods

### Study population

We used data from the UK Biobank (UKB), a population-based prospective cohort study of 502,419 UK residents aged 40-69 years recruited between 2006-2010 from 22 assessment centers across the UK.^31^ A wide range of phenotyping assessments, biochemical assays, genome-wide genotyping, and ongoing longitudinal follow-up data is available for the vast majority of study participants. For the purposes of the current analyses, we restricted our sample to 46% of the study participants (*n*=231,336) with detailed linked electronic medical records from their primary care general practitioners (GP). These primary care data include medication prescriptions for a time period ranging from as early as 1978 until 2019, thus allowing a detailed assessment of duration and dose of statin intake both before and after baseline assessments. We excluded 15,137 individuals with missing genetic data and 155 individuals with a history of ICH at baseline (defined as presence of a prevalent diagnosis code in the UKB assessment center, **Fig. 1**).

**Figure 1.**
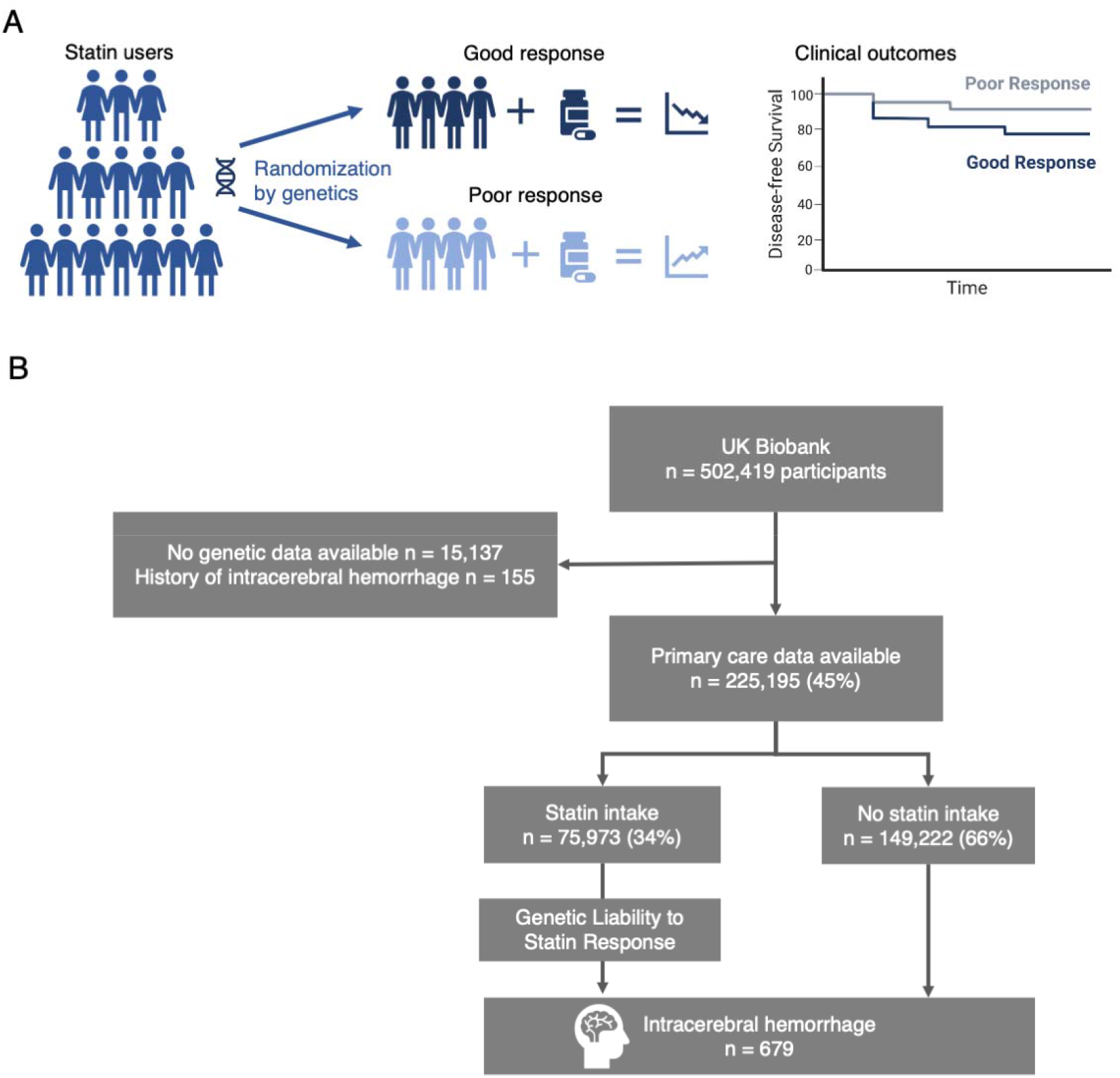
Study concept and study population. (**A**) Individuals respond differently to statins according to their genetic profile.^37^ Assuming that there is no selection pressure, drug response is assorted randomly within a population and can be used to explore causal effects of the drug on outcomes using observational data. (**B**) Flowchart of the study participants. Individuals without genetic data or history of intracerebral hemorrhage at baseline were excluded.

The UKB has institutional review board approval from the Northwest Multi□Center Research Ethics Committee (Manchester, UK). All participants provided written informed consent. We accessed the data following approval of an application by the UKB Ethics and Governance Council (Application No. 36993).

### Preparation of the UK Biobank primary care data

A detailed description of the primary care data including limitations is available from the UK Biobank (https://biobank.ndph.ox.ac.uk/showcase/showcase/docs/primary_care_data.pdf). We extracted data on-statin prescriptions from the GP prescriptions table by using international nonproprietary (INN) names, former and current trade names, Dictionary of Medicines and Devices (DM+D) and British National Formulary (BNF) codes of all available ever approved statins. The list of trade names was obtained by queries to the DM+D product search browser (https://services.nhsbsa.nhs.uk/dmd-browser/search). The list of terms and codes used for searching the prescription data is given in **Supplementary Table S1**. We used only prescriptions for dates after 1990, which accounted for the vast majority (99.99%) of the data and also coincided with the time period when wide usage of statins began.^32^ The dosages were extracted from the prescription texts by using regular expressions with subsequent manual curation of entries (**Supplementary Table S1**). As quality control measures, we removed duplicate prescriptions for the same data per individual and in case of multiple different statins prescribed for the same date (0.4%), we randomly removed all but one prescription. We defined statin users as individuals who had at least one statin prescription between 1990 and end of follow-up. For obtaining statin exposure metrics, we harmonized the dosages of different statins based on comparison factors from trials evaluating statin efficacy.^3,33-35^ We averaged the slightly different comparison factors to assign potency equivalency factors of 20, 2, 0.5, 0.25, 0.125, for cerivastatin, rosuvastatin, simvastatin, pravastatin, and fluvastatin, respectively, using atorvastatin as the reference statin.^3,33-35^ We then calculated a cumulative statin exposure in units of 10mg atorvastatin/5 years.

Beyond statin prescriptions, we extracted LDL cholesterol measurements from the primary care data. We gathered all available codes for LDL measurements by an automatic search for “LDL” in the lookup table of GP codes and following curation of the code list, we used these codes to extract LDL measurements from the GP clinical event records table (**Supplementary Table S2**). Only two of the codes specified that LDL was calculated (14.8% of the values obtained), for the rest of the codes it was unknown whether LDL was directly measured or calculated. Information on measurement assay was unavailable for all codes. Values were converted from mmol/l to mg/dl by multiplying with the factor 38.67.^36^ As quality control measures, we removed entries with measurement date before 1990 or after 2022 (0.003%), entries with empty values (3.6%), entries with values over 300 mg/dl (10.3 mmol/l) or less than 10 mg/dl (0.26 mmol/l) to account for miscoding in mg/dl (0.04%), and duplicate entries.

### Polygenic score for estimating on-statin LDL response

We used data from the Genomic Investigation of Statin Therapy (GIST) Consortium, a two-stage genome-wide association study (GWAS) for on-statin LDL cholesterol response among 40,914 statin-treated subjects of Eureopean ancestry (30,246 from 10 randomized controlled trials and 10,668 from 11 observational studies),^37^ to construct a polygenic score of LDL-lowering following statin intake. There was no participant overlap between those studies and the UK Biobank. Following a previously described approach, we used a set of 35 single nucleotide polymorphisms (SNPs), selected based on associations with on-statin LDL lowering at *p*<5×10^−5^ and clumped at r^2^<0.001 based on the European reference panel of the 1000 Genomes.^38^ We then calculated a genetic score with the imputed genotype data of UK Biobank. To confirm that the observed effects were specifically due to genetically predicted on-statin LDL and not off-statin LDL, in sensitivity analyses we tested the association of each of the 35 SNPs included in the score and LDL levels measured at the UK Biobank baseline assessment among never-statin users (linear regression models adjustments for age, sex, principal components 1-10 of population structure, kinship, and genotyping assay) and removed the SNPs that associated with off-statin LDL at *p*<0.0014 (0.05/35 according to Bonferroni). All SNPs for the genetic and the alternative score are provided in **Supplementary Table S3**.

### Validation of statin response genetic scores on LDL trajectories

To test the relevance assumption of MR, we aimed to confirm the effect of the employed genetic score on on-statin LDL levels by exploring associations with longitudinal LDL level changes in the primary care data among statin users. Only participants with at least one LDL measurement before and one measurement after their first recorded statin prescription (off- and on-statin LDL) were included in this analysis (*n=*40,633, 53% of statin users). To account for multiple LDL values per participant over time, we used a mixed model clustered by participant with LDL levels as the outcome and the genetic score, time, and their interaction as the exposure. The modes were further adjusted for age, sex, statin equivalency dose, PC1-10, race, kinship, and genotyping assay.

### Influence of the genetic scores on baseline LDL and lipid particle metabolites

To explore whether a higher genetic score for on-statin LDL-lowering mimics an exposure to higher statin intake, we compared associations of a higher score and a higher statin dose with the entire spectrum of 228 lipid particle metabolites among statin users, as measured by nuclear magnetic resonance (NMR) at baseline. We constructed linear regression models with each metabolite as outcome and the genetic scores or statin equivalent dose as exposure. The models were further adjusted for age, sex, statin equivalency dose, PC1-10, race, kinship, and genotyping assay. We corrected for multiple hypothesis testing with the Bonferroni method setting a significance threshold at 0.05/228. Correlations in the derived estimates for the genetic scores or statin equivalent dose across the lipid traits were tested with the Pearson’s correlation.

### Outcome ascertainment

UKB participants’ records have been linked with inpatient hospital episode statistics (HES), primary care data, and death registry for longitudinal follow-up. Incident ICH events were captured by the diagnostic algorithm for stroke in the UKB (https://biobank.ndph.ox.ac.uk/showcase/ukb/docs/alg_outcome_stroke.pdf) up to December 2018. For events occurring thereafter and up to the end of follow-up (June 2020), we used the respective ICD-9 and ICD-10 codes in HES or death registry data (ICD-9 431.X and ICD-10 I61). As positive controls, we also tested associations of the genetic scores with incident myocardial infarction (MI) and peripheral artery disease (PAD), which were defined on the basis of the following ICD-10 codes: I21.X, I22.X, I23.X, I24.1, I25.2 (for MI) and I70.0, I70.00, I70.01, I70.2, I70.20, I70.21, I70.8, I70.80, I70.9, I70.90, I73.8, I73.9 (for PAD).

### Effect of the genetic score on ICH and cardiovascular endpoints

To explore the effects of on-statin genetically predicted LDL response on risk for incident ICH, we used Cox proportional hazard models adjusted for previously published risk factors for ICH^1,2^ and genetic covariates: age, sex, BMI, smoking status, history of diabetes, systolic blood pressure, cumulative statin dose exposure, use of anticoagulation and antiplatelet drugs at baseline, PC1-10, race, kinship, and genotyping assay. We constructed separate models with and without LDL. As positive controls, we explored associations between on-statin genetically predicted LDL response and risk for MI and PAD using similar Cox models but without adjusting for antiplatelet and anticoagulation intake. To test the independence and exclusion restriction assumptions of MR and exclude the possibility that any associations are driven by pleiotropic effects of the score independent of on-statin LDL-lowering, we tested the same associations among non-statin users. Because statin users are, due to indication bias, at higher risk for MI and PAD, the selection of the population based on statin use could have introduced collider bias. To address this issue, in a sensitivity analysis we used inverse probability weighting to confirm our findings for MI and PAD. Specifically, in the full UK Biobank cohort, we constructed a linear regression model with statin use as outcome and age, sex, BMI, smoking status, hypertension, systolic blood pressure, history of diabetes, intake of diabetes drugs, hypercholesterolemia, LDL, history of MI, stroke or PAD, and the genetic score as covariates. For statin users, we then used the inverse of the fitted values of that model as weights in the respective Cox models to account for the probability of statin prescription in an individual.

### Software used

For SNP extraction, genetic score calculation, SNP association tests, and relationship inference we used PLINK, bcftools, and KING.^39-42^ For data extraction, curation, preparation, and figure generation, we used RStudio 2021.09.0 with R version 4.1.1 on Mac OS X (aarch64-apple-darwin20) with the packages coxphw, data.table, dplyr, FSA, ggplot2, gmodels, lmerTest, lme4, PheWAS, readr, readxl, stringr, survival, survivalAnalysis, survminer, tidyr, and writexl.^43^ The analysis plan followed the STROBE-MR statement for the usage of Mendelian Randomization in observational studies.^44^

### Data availability

The data that support the findings of this study are available from the UK Biobank upon submission of a research proposal. The summary statistics of the GWAS for on-statin LDL response used to create the tested polygenic risk score are publicly available.^31,37^

## Results

### Baseline characteristics

A total of 225,195 of the UKB participants had available genetic, primary care, and outcome data, as well as no history of ICH at baseline and were thus eligible for inclusion in the analysis (**Fig. 1**). Baseline characteristics and outcome data of participants included in the analyses are presented in **Table 1**. A total of 4,151,471 statin prescriptions for 6 statin agents were extracted from the primary care data. The majority of the prescriptions referred to simvastatin (60.8%) or atorvastatin (30.4%) and the number of statin prescriptions increased over time. We extracted at least one statin prescription for 75,973 of the participants (33.7%) with available primary care data. 41% of statin users had prescriptions for two or more different drugs at different time points indicating a medication switch. The distributions of statin prescriptions over time, age, dose, and presence of vascular risk factors are depicted in **Fig. 2** and the detailed distribution of different statins is shown in **Supplementary Table S3**. Of all prescriptions, 17.1% accounted for a low (≤10mg), 30.5% for a medium (>10 and ≤20mg), and 52.3% for a high (>20mg) atorvastatin equivalency dose with proportion of individuals prescribed a statin and statin dose increasing with age. Similarly, statin use and higher statin doses were more common among individuals with more vascular risk factors (active smoking, diabetes, hypertension, hypercholesterolemia, prevalence of MI, stroke, or PAD, or age > 65 years) (**Fig. 2**).

**Table 1.** Participant characteristics.

| **Baseline characteristics** | **All** | **Statin users** | **Statin non-users** |
| --- | --- | --- | --- |
| **All, n** | 225,195 | 75,973 | 149,222 |
| **Female, n (%)** | 122,793 (54.5) | 32,491 (42.8) | 90,302 (60.5) |
| **European genetic ancestry, n (%)** | 201,856 (89.7) | 67,968 (89.5) | 133,888 (89.8) |
| **Age, years, mean ± SD** | 56.5 ± 8.1 | 60.4 ± 6.6 | 54.6 ± 8.0 |
| **BMI, kg/m<sup>2</sup>, mean ± SD** | 27.5 ± 4.8 | 28.9 ± 4.9 | 26.8 ± 4.6 |
| **Ever Smokers, n (%)** | 101,098 (44.9) | 40,382 (53.4) | 60,717 (40.8) |
| **Former Smoker, n (%)** | 77,469 (34.4) | 30,884 (40.9) | 46,585 (31.3) |
| **Current Smoker, n (%)** | 23,630 (10.5) | 9,498 (12.6) | 14,132 (9.5) |
| **Hypertension, n (%)** | 60,036 (26.7) | 36,202 (47.7) | 23,834 (16.0) |
| **Systolic blood pressure, mmHg, mean ± SD** | 141.3 ± 20.6 | 150.0 ± 20.4 | 136.9 ± 19.3 |
| **Diastolic blood pressure, mmHg, mean ± SD** | 84.5 ± 11.3 | 88.1 ± 11.3 | 82.7 ± 10.9 |
| **Hypercholesterolemia, n (%)** | 27,573 (12.2) | 24,708 (32.5) | 2,865 (1.9) |
| **LDL, mg/dl, mean ± SD** | 137.8 ± 33.9 | 134.4 ± 39.9 | 139.5 ± 30.2 |
| **HDL, mg/dl, mean ± SD** | 56.3 ± 14.7 | 52.6 ± 13.9 | 58.0 ± 14.7 |
| **Diabetes, n (%)** | 11,370 (5.0) | 9,798 (12.9) | 1,527 (1.1) |
| **Antiplatelet use, n (%)** | 33,056 (14.6) | 23,971 (31.6) | 9,085 (6.1) |
| **Anticoagulant use, n (%)** | 2,541 (1.1) | 1,698 (2.2) | 843 (0.6) |
| **Incidence of outcomes after enrollment** |  |  |  |
| **Intracerebral hemorrhage, n (%)** | 679 (0.3) | 383 (0.5) | 296 (0.2) |
| **Myocardial infarction, n (%)** | 12,355 (5.5) | 8,756 (13.0) | 3,599 (2.4) |
| **Peripheral artery disease, n (%)** | 1,711 (0.8) | 1,378 (1.8) | 333 (0.2) |
| **Ischemic stroke, n (%)** | 2742 (1.2) | 1,973 (2.6) | 769 (0.5) |

Abbreviations: LDL, low-density lipoprotein; HDL, high-density lipoprotein; SD: standard deviation.

**Figure 2.**
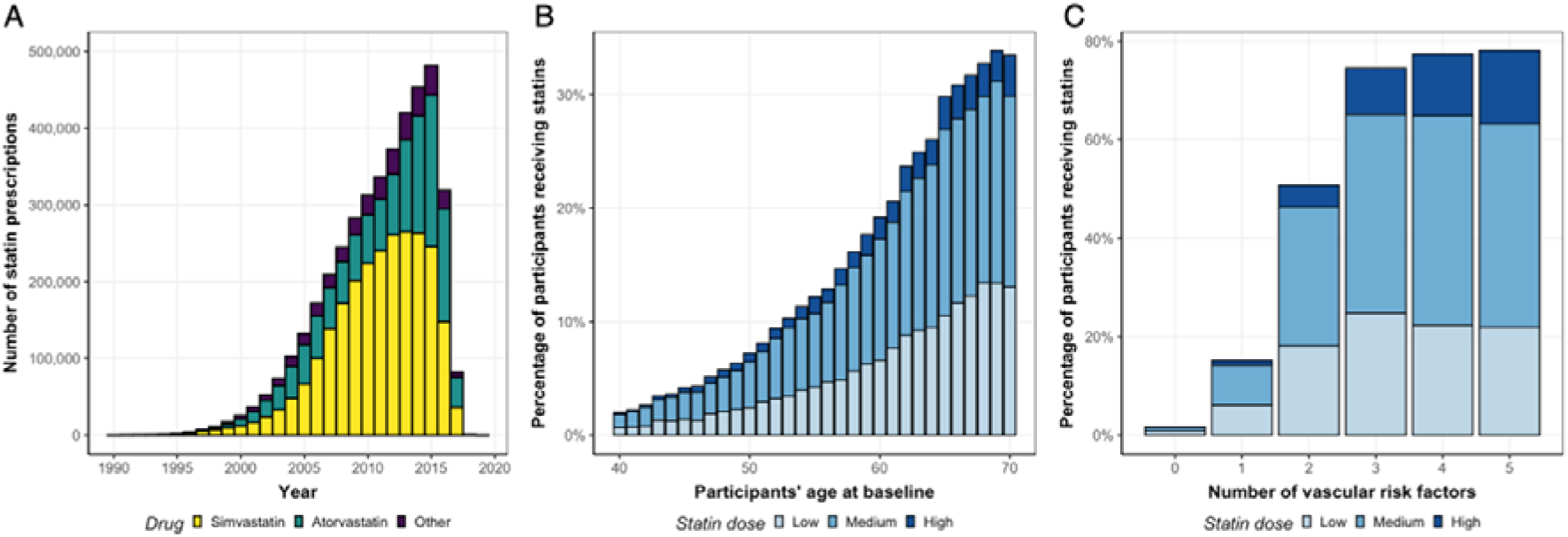
Statin prescriptions in the UK Biobank primary care data. **(A**) Number of statin prescriptions per year among 75,973 statin users. (**B**) Percentage of participant’s receiving statins per participant’s age at baseline. (**C**) Percentage of participants at baseline receiving statins per number of vascular risk factors (diabetes mellitus, hypercholesterolemia, hypertension, active smoking, age > 65 years). Statin doses in B and C were divided in low (≤ 10mg), medium (> 10mg and ≤ 20mg), and high (> 20mg) Atorvastatin equivalent dose. Estimated drug potencies were used to harmonize all drug doses to Atorvastatin equivalent doses (see Methods).

### Genetic score for on-statin LDL and LDL trajectories in primary care data

To validate the genetic scores for on-statin LDL lowering in the UKB, we extracted LDL measurements recorded in the primary care data. A total of 46,909 participants (62% of the total statin users) had at least one LDL value before and one after their first statin prescription. There were on average 8.1 ± 5.1 measurements spanning a total of 27.4 years (8.7 ± 4.2 years between first and last measurement). The mean pre-statin LDL was 147.7 ± 38 mg/dl and the mean post-statin LDL 133.0 ± 42.7 mg/dl. The mean LDL decreased significantly over time (−3.45 mg/dl per year, 95% CI: [-3.47, -3.42]) among statin users. In a mixed linear model adjusting for age and sex, the genetic score and statin dose both had significant effects on absolute LDL levels (−2.3 mg/dl for each SD increase of genetic score, 95% CI: [-2.59, -2.00], **Fig. 3A**, and -18.8 mg/dl for each SD of statin dose, 95% CI [-18.91, -18.66], **Fig. 3B, Supplementary Table S4**). Importantly, there was a significant interaction of the genetic score with time implying a more rapid on-statin LDL decrease among participants with a higher genetic score (−0.05 mg/dl per year for one SD of the genetic score, 95% CI [-0.07, -0.02]).

**Figure 3.**
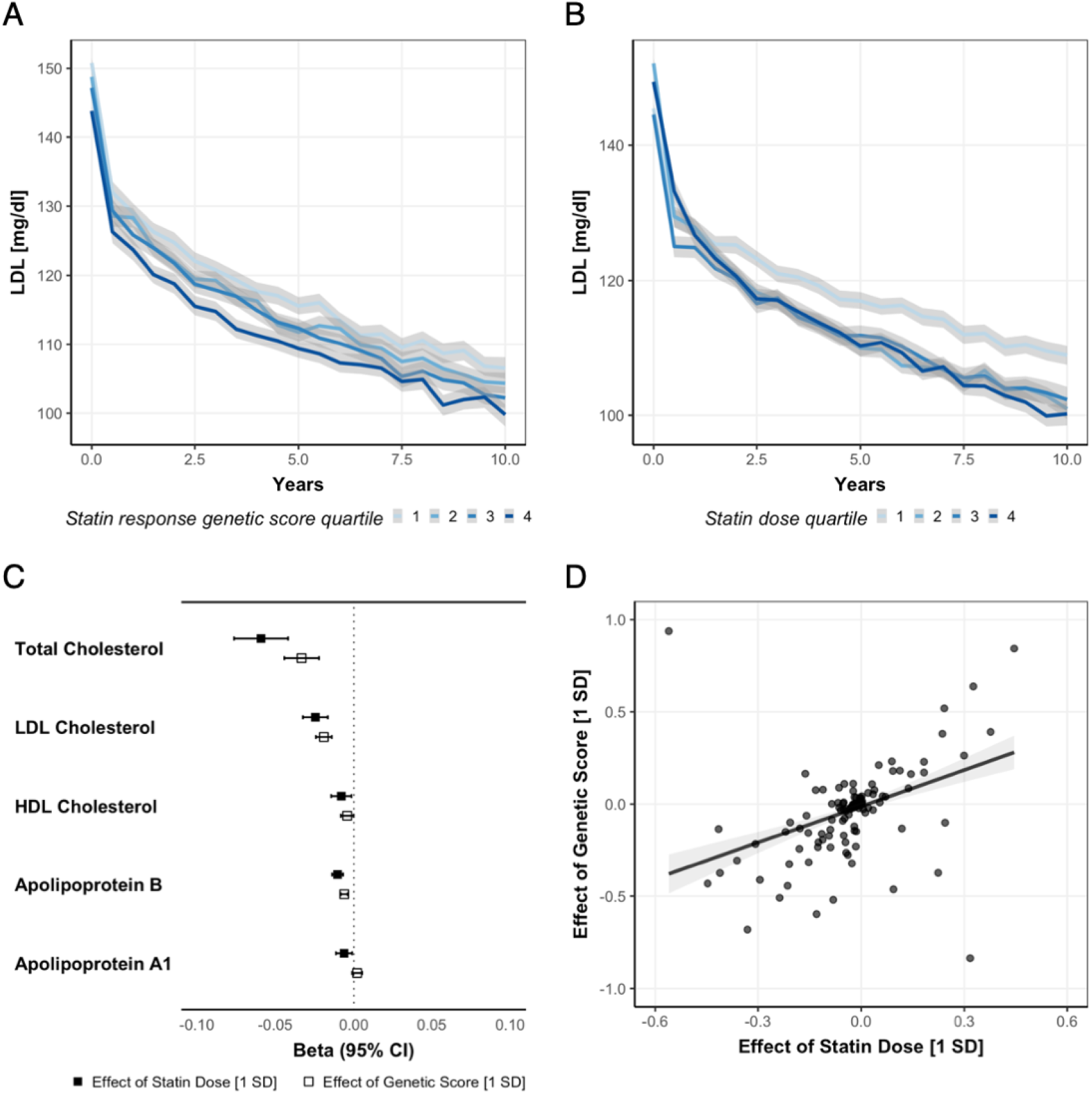
Effect of the genetic score and statin dose on on-statin LDL levels (A, B) and lipid metabolites (C, D) among statin users. LDL trajectories derived from the primary care data of the UK Biobank separated (**A**) by quartiles of the genetic score for LDL response after statin intake and (**B**) by mean statin dose over lifetime in 46,909 individuals with statin intake and at least one LDL measurement before the first statin prescription. (**C**) Estimate and 95% confidence intervals of the effect of statin dose and the genetic score for statin response on total, LDL, and HDL cholesterol (mmol/l) and on apolipoprotein B and A1 (g/l) derived from NMR among statin users. (**D**) Comparison of the effect size of the genetic score for statin response (1 SD increment) vs the statin dose (1 SD increment) on lipidomic metabolites among statin users. Each point represents the effect sizes for one of 228 lipidomic particles. Correlation coefficient *r* = 0.65. The results from C and D are derived from linear regression models adjusted for age, sex, PC1-10, race, kinship, and genetic assay.

Because we found the genetic score to be also associated with absolute off-statin LDL levels among non-users (−3.0 mg/dl for each SD increase of genetic score, 95% CI: [-3.3, -2.8]), we tested the effect of each SNP and found four of them to be significantly associated with off-statin LDL levels (**Supplementary Table S5**). Thus, in a sensitivity analysis, we constructed an alternative genetic score with the remaining 31 SNPs that was no longer associated with off-statin LDL levels among non-users (*p*>0.05), but was associated with significant on-statin LDL-lowering among statin users (−0.03 mg/dl per year per one SD, 95% CI [-0.05, -0.01]).

### Influence of the genetic scores on lipid particle metabolites

Next, to explore whether a higher genetic score for on-statin LDL lowering mimics an exposure to higher statin intake at a metabolomic level, we investigated its effects on cholesterol measurements, as well as lipid particle metabolites, as assessed by standardized methodologies at baseline among statin users. The comparisons of the effect sizes of statin intake on NMR-assessed lipid particle metabolites are presented in **Fig. 3C** and **3D**, respectively. Of the 228 lipid particle metabolites analyzed, the genetic score was significantly (Bonferroni-adjusted *p*<0.05) associated with 161 and a higher statin dose with 97 (**Supplementary Table S6**). There was a correlation between the effect sizes of the genetic score and statin dose (*r* = 0.52, *p*<0.001).

### Genetically predicted on-statin LDL-lowering and risk of incident ICH

Following the validation of the genetic score as a proxy of on-statin LDL-lowering, we next tested associations with the risk of incident ICH among statin users (**Fig. 4**). There were 679 incident ICH over an observation period of 2,514,994 person-years, yielding an incidence of 27 per 100,000 person years. Over a mean follow-up of 11 years, 383 statin users developed ICH. In Cox proportional hazard models, higher genetic scores for on-statin LDL-lowering were associated with a higher risk of incident ICH among statin users (HR 1.15, 95% CI [1.04, 1.27] for one SD difference). Sensitivity analyses confirmed robustness of the findings among unrelated individuals (kinship coefficient <0.0884, *n*=70,220; HR per-SD increment 1.17, 95% CI [1.06, 1.30]) as well as with the alternative genetic score not influencing off-statin LDL levels (HR per-SD increment 1.13, 95% CI [1.02, 1.24]). As positive controls, we tested the effects of genetically predicted on-statin LDL-lowering on MI and PAD. After adjustment for cardiovascular risk factors, LDL levels, and statin dose, we found significant associations of a higher genetic score with a lower risk of both incident myocardial infarction (HR per-SD increment 0.98 95% CI [0.96, 0.99]) and PAD (HR per-SD increment 0.92 95% CI [0.86, 0.98]). To reduce the risk for collider bias, we calculated models weighted for the inverse probability of being prescribed a statin, which yielded effect estimates in the same direction for both MI (HR per-SD increment 0.97 95% CI [0.94, 0.99]) and PAD (HR per-SD increment 0.84 95% CI [0.72, 0.99]). Finally, to minimize the possibility that the observed effects are the result of pleiotropy on traits other than on-statin LDL, we also tested the same associations among statin non-users and found no significant effects on ICH, MI, or PAD (all *p*>0.05).

**Figure 4.**
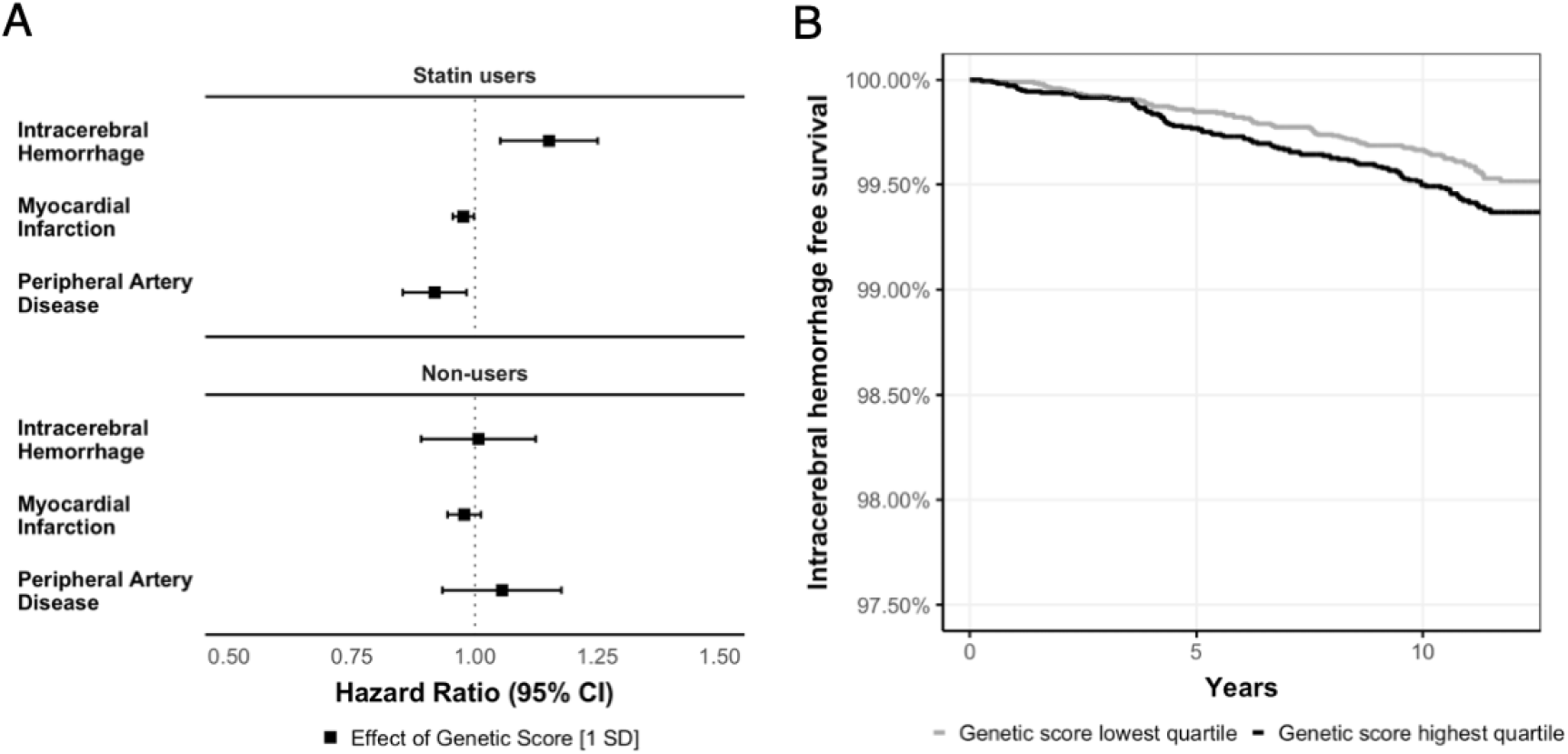
Effect of on-statin genetically predicted LDL response on study outcomes. **(A**) Hazard ratios of statin dose and the genetic scores for statin response among statin users and non-users on intracerebral hemorrhage, myocardial infarction, and peripheral artery disease. The results are derived from Cox proportional hazard models adjusted for age, sex, BMI, smoking status, history of diabetes, systolic blood pressure, cumulative statin dose exposure, PC1-10, race, kinship, and genotyping assay; use of anticoagulation and antiplatelet drugs at baseline in models for intracerebral hemorrhage. (**B**) Kaplan-Meier curves for survival intracerebral hemorrhage in statin users.

## Discussion

In this study, we used genetic data to stratify statin users by their genetically predicted response to statins and investigated their risk of incident ICH risk. We leveraged data from a GWAS of on-statin LDL lowering from 40,000 statin-treated individuals (75% clinical trial participants) as well as biochemical, lipidomic, and primary care data from 225,000 individuals from a population-based study. We found that a higher genetically predicted LDL response to statins associated with steeper LDL lowering, a similar lipidomic signature as high-dose statin use, and a lower risk of atherosclerotic cardiovascular outcomes. In addition, this higher genetically predicted LDL response to statins was associated with a higher risk of ICH among statin users only. There was no such association among individuals who were not taking statins. Our results support a causal effect of more aggressive LDL-lowering with statins on risk of ICH and highlight the utility of modeling drug response in addition to dose in examining putative causal associations between biomarkers and outcomes.

Our study extends previous findings from genetic^26,29^ and observational analyses,^8-13^ providing evidence that beyond lifetime variation in LDL levels, genetic variation in statin-induced LDL-lowering also influences ICH risk. This result agrees with post hoc analyses of clinical trials supporting a higher risk for hemorrhagic stroke among participants prescribed a high-intensity statin dose.^18^ The mechanisms underlying this observation remain poorly understood. It has been speculated that cholesterol is important for vessel integrity, but to date no experimental study has provided evidence for a mechanism connecting low cholesterol levels to vessel damage or loss of vessel structural integrity.^45^ As demonstrated in our analyses but also in previous work,^46^ statins influence a wide range of lipoprotein particles beyond LDL and thus revealing the main driver of their association with ICH remains a key challenge. Aggressive LDL-lowering by PCSK9-inhibitors does not increase ICH incidence even in high-risk patients with prior ischemic or hemorrhagic stroke, indicating that LDL might not be the sole driver.^17^ Because of the widespread lipidomic effect of the genetic score we used, it is not possible from our current analyses to make inferences about which particle class is the causal mediator of this association.

By leveraging genetic determinants of response to statin intake, we were able to randomize statin users at the beginning of drug intake, as the prescribing physician is blinded to the genetic variation in statin response. In contrast, when using genetic variants for off-statin LDL or HDL levels in conventional MR approaches,^26-29^ randomization is performed at conception and leads to lifelong variations in lipid levels. As such, conventional MR studies have captured lifelong genetically predicted LDL levels and are thus limited in making any inferences about the causal effects of a particular drug prescribed over a shorter timeframe. Our approach overcomes this limitation, facilitating causal inference of the impact of statin intake on ICH using solely observational data. This application could be implemented in other settings as well, and demonstrates the latent utility of additional efforts to develop polygenic predictors of drug response in pharmacogenomic research.^30^

From a methodological perspective, our study also demonstrates the utility of using real-world primary care data for assessing longitudinal trajectories of clinical and biochemical assessments and medication use. Although real-world data are noisier and less standardized than data usually obtained for research purposes, they retain utility to assess drug safety and side effects, inform clinical trial design, and compare drug effectiveness.^47^ Leveraging the longitudinal drug prescription and LDL measurement data from primary care data, we were able to track statin prescription and response over a timeframe extending from several years before inclusion of the participants to the study to the end of their follow-up in the UK Biobank. Using data from the rising number of GWAS for drug response,^48,49^ future studies could explore in the primary care data from the UK Biobank associations of drug intake with multiple endpoints. This could allow the detection of previously unreported adverse effects, for which trials are often underpowered,^50^ or the investigation of the potential of repurposing opportunities.

Our study has additional specific methodological strengths. Using data from 225,000 participants, including 75,000 statin users and 700 ICH events, we were sufficiently powered to detect meaningful changes in ICH risk by genetically predicted on-statin response. The phenotypic depth of the UK Biobank dataset allowed us to validate the effects of the genetic score statin response on LDL trajectories, lipidomic traits, and atherosclerotic endpoints. Furthermore, we have introduced novel and innovative approaches to leverage GWAS for drug response in large-scale longitudinal population-based datasets. By aggregating data from >4 million drug prescriptions, we were able to precisely phenotype drug intake at an individual level and thus control for statin dose in our outcome models.

Our approach also has limitations. First, the constructed genetic score was associated not only with on-statin LDL lowering but also with off-statin baseline LDL levels. To address this limitation, we introduced an alternative genetic score which was only associated with LDL lowering after statin intake and used that for sensitivity analyses confirming our findings. However, residual confounding due to subthreshold effects of the variants on baseline LDL levels cannot be excluded. Second, we observed a lower incidence of ICH in our study population (27 per 100,000 person-years), as compared to the age-standardized world-wide rate of 42 per 100,000 person-years.^1^ This is possibly related to the healthier profile of the UK Biobank population as compared with the general population and necessitates a cautious interpretation of the findings.^51^ Third, our study was performed in mainly people of European ancestry and therefore our results cannot be generalized to other populations. Fourth, actual drug intake might also be influenced by poor adherence, which has not been included in our models. Fifth, statins were first introduced in 1988 and prescriptions rose since then, but it was not until 1995 that more than 90% of the primary care practices in the UK were fully computerized,^52^ leading to a possible underestimation of the cumulative statin doses that we used in our models. Finally, because atherosclerotic cardiovascular disease prevention is the main indication for statins, limiting our cohort to statin users might have introduced collider bias for the atherosclerotic endpoints.^53^ While we addressed this issue by applying inverse probability weighted models, some relevant bias towards the null might still be present in the measured effect sizes.

In conclusion, we found that higher genetically predicted on-statin LDL response mimics exposure to higher statin doses and increases risk for ICH. These results imply that more aggressive statin-induced LDL lowering might increase risk of ICH and should be balanced against statin benefits in trials of intensive statin treatment. More broadly, our results demonstrate the utility of leveraging genetic data of drug response as a novel method of investigating side effects and repurposing opportunities of specific drugs with observational data.

## Data Availability

The data that support the findings of this study are available from the UK Biobank and the GWAS for on-statin LDL.

## Abbreviations

GP: General Practitioner
GWAS: Genome-wide Association Study
HDL: High-density Lipoprotein
HR: Hazard Ratio
ICH: Intracerebral Hemorrhage
LDL: Low-density lipoprotein
MI: Myocardial Infarction
MR: Mendelian Randomization
PAD: Peripheral Artery Disease
SD: Standard Deviation
SNP: Single Nucleotide Polymorphism
UK: United Kingdom
UKB: UK Biobank

## Acknowledgements

This research has been conducted using data from UK Biobank, a major biomedical database (www.ukbiobank.ac.uk) under the project ID 36993 “Identification of genetic components underlying cerebrovascular diseases and their related outcomes”.

## Supplementary material

Supplementary material is available at *Brain* online.

